# The ILANA study: a paradigm shift in ensuring equity of clinical implementation in HIV research

**DOI:** 10.1101/2022.11.30.22282915

**Authors:** Hamzah Z Farooq, Vanessa Apea, Bakita Kasadha, Sadna Ullah, Gill Hilton-Smith, Amber Haley, Jenny Scherzer, James Hand, Sara Paparini, Rachel Phillips, Chloe Orkin

**Author notes:** **Corresponding Author:** Professor Chloe M Orkin, Email -, **Address:** SHARE Collaborative, Blizard Institute, Queen Mary University of London, 4 Newark St, London, E1 2AT, UK. **Ethical approval and consent to participate:** Study was reviewed and approved by Research Ethics Committee and Health Research Authority. Reference: 22/PR/0318. **Trial registration:** Clinicaltrials.gov ref: NCT05294159. **Consent for publication:** All authors gave their consent for publication. **Availability of data and materials:** All the data for this study will be made available upon reasonable request to the corresponding author. **Funding:** ViiV Healthcare Ltd, research grant to Queen Mary University of London.

## Abstract

**Introduction:** Cabotegravir and Rilpivirine (CAB+RPV-LA) is recommended as a treatment for HIV-1 allowing people living with HIV to receive two-monthly injectable treatment, rather than daily pills. Providing injectable therapy in a system designed to provide and manage patients on oral treatments poses logistical challenges namely how resources are used to accommodate patient preference within constrained health economies with capacity limitations. In this pragmatic multi-centre study, we aim to understand the implementation of CAB-RPV-LA administration in two settings via mixed methods to explore perspectives of participants and the clinical team delivering CAB+RPV-LA.

**Methods and Analysis:** Women, racially minoritised people and older people are chronically under-represented in HIV clinical trials so the ILANA trial has set recruitment caps to ensure recruitment of 50% women, 50% ethnically-diverse people and 30% over 50 years of age to include a more representative study population. Utilising a mixed-methods approach, the primary objective is to identify and evaluate the critical implementation strategies for CAB+RPV-LA in both hospital and community settings. Secondary objectives include evaluating feasibility and acceptability of CAB+RPV-LA administration at UK clinics and community settings from the perspective of HIV care providers, nurses, and representatives at community sites, evaluating barriers to implementation, the utility of implementation strategies, and adherence.

**Ethics and Dissemination:** Ethical approval has been obtained from the Health Research Authority Research Ethics Committee (REC reference: 22/PR/0318).

The dissemination strategy has been formulated with the SHARE Collaborative Community Advisory Board in order to maximise the impact of this work on clinical care and policy. This strategy draws upon and leverages existing resources within the participating organisations, such as their academic infrastructure, professional relationships and community networks fully. The strategy will particularly harness the Public Engagement Team and press office to support dissemination of findings.

**Registration Number:** ClinicalTrials.gov Identifier: NCT05294159

**Strengths and Limitations of the Study:**

1. This trial employs an anti-racist, anti-sexist, anti-ageist approach to protocol design, building equitable recruitment into the fabric of the protocol.
2. This is the first implementation study to evaluate delivery of long-acting injectable HIV antiretrovirals (LAIs) in both community and clinic settings and the first UK-based trial of LAI to evaluate routine clinical practice within the National Health Service (NHS).
3. The trial also contains a mixed-methods sub-study exploring reasons for trial non-participation.
4. Trial sites are all large, urban centres. Further studies of implementation of LAIs in smaller and rural settings will be needed.
5. The small sample size and specific targets for women and racially-minoritised groups aims to be representative of people living with HIV in the UK, but may not be representative of all people choosing the option of injectable medication.

## Introduction

HIV has been one of the greatest infectious challenges of our time. According to the World Health Organization, 38-million people were living with HIV people living with HIV worldwide in 2019, 51% are women and over 50% are racially-minoritised ^(1-3)^. Anti-retroviral Therapy (ART) not only prevents progressive, fatal illness but affords a near-normal life expectancy to people living with HIV who are treated early ^(4)^. However, for those who find oral therapy difficult or impossible to take, it remains a fatal illness.

Many studies have demonstrated demand and interest in long-acting therapies in people living with HIV and are regarded as a welcome development by many ^(5-7)^. Long-acting injectable Cabotegravir and Rilpivirine (CAB+RPV-LA) has been shown to be effective and safe as a complete regimen delivered either monthly or two-monthly in three large phase III clinical trials ^(5-7)^. Participants on CAB+RPV-LA overwhelmingly preferred it to daily oral therapy and had statistically significantly higher satisfaction and acceptance scores according to validated patient reported outcome measures ^(8).^ CAB+RPV-LA therapy has been shown to be highly acceptable^9,10^ and removes the burden of daily oral therapy for people living with HIV. This can contribute to achieving viral suppression for those who, for multiple complex reasons, can’t or won’t take oral therapy.

Current HIV clinical services in the UK are designed to provide care to people on oral therapy who are generally monitored six-monthly. Providing injectable therapy within this resource-limited system poses logistical challenges, namely how resources are used to accommodate anticipated patient preference for long-acting treatment ^(9)^. Common challenges to implementation (such as prioritization of patient populations for preferred use, clinic infrastructure requirements, steady supply chains, provider and patient training) require conducting rigorous implementation science research to ensure this treatment reaches patients who would most benefit from it ^(10)^.

The NHS provides free ARTs to all, regardless of immigration status. CAB+RPV-LA is particularly important for those who experience pill fatigue, HIV-related stigma and have fears of inadvertent disclosure. This is highly relevant to the ethnically diverse population of people living with HIV in the UK, many of whom come from marginalised and minoritised communities in which HIV-related stigma is common and treatment outcomes are the poorest ^(11)^.

Women, racially-minoritised people and older people are chronically under-represented in HIV clinical trials ^(12, 13)^, yet disproportionately affected by HIV ^(12, 14)^. Fifty-one percent of people living with HIV are women ^(15)^. Fifty-seven percent of heterosexuals people living with HIV in the UK are of Black African heritage ^(16)^ and 14% of men-who-have-sex-with men (MSM) living with HIV are from racially-minoritised backgrounds ^(16)^.

Achieving equity of care for women and racially-minoritised people living with HIV requires a bold and inclusive approach to research ^(17)^ which is why we have developed this protocol for implementing LAIs in multiple settings, with a focus on under-represented populations. The ILANA study aims at exploring patients’ experiences and perceptions of the long-acting injectable CAB+RPV and its facilitation in the NHS. An anti-racist, anti-sexist, anti-ageist approach to recruitment is written into the protocol and stipulates that male participants and White participants will be capped at 50%, with a cap at 70% for patients younger than 50 years of age.

In this pragmatic real-world trial, LA CAB+RPV will be delivered in the clinics for the first six-months and each site will identify the most pragmatic community setting in which to deliver CAB+RPV-LA during the second six-months as an additional option alongside those who choose to continue receiving care in hospital clinics. Consequently, this study explores implementation of a standard-of-care treatment in two types of setting. The study includes a mixed-methods sub-study (NO-LANA) exploring reasons for non-participation in individuals who opted not to take part in the main ILANA study.

## Aims and objectives

This research aims to understand the implementation of CAB-RPV-LA administration in two settings – the HIV clinic and in the community (eg, pharmacy, home, community organisation), via a mixed methods approach exploring perspectives of participants and the clinical team delivering CAB+RPV-LA in both settings.

The objectives are as follows:

### Primary objective

- To evaluate feasibility of CAB+RPV-LA administration at NHS HIV clinics and community settings in people living with HIV who will receive CAB+RPV as part of their routine clinical care.

### Secondary objectives

1. To evaluate the feasibility and acceptability of CAB+RPV-LA administration at six clinics and community settings in England for HIV care providers, and nurses
2. To evaluate the utility of and fidelity to Facilitation Calls and Blueprints (Standard Operating Procedure) for the Community Nurse or Clinic Nurse
3. To describe barriers and facilitators to implementation of CAB+RPV-LA from the perspective of HIV clinic staff, Community Nurses, Clinic Nurses and participants living with HIV
4. Describe the preferences of people living with HIV for the setting they receive injections and reasons for their choice
5. Describe any change in treatment satisfaction scores and tolerability and acceptance of injections, over time and by setting
6. To describe adherence to the dosing window

### Tertiary objectives

- To describe proportion virologically suppressed (HIV viral load < 50 c/ML) and the safety of CAB+RPV-LA
- To understand reasons for non-participation in the study

### Research questions

- What are the experiences and perceptions of patients and healthcare-providers of the acceptability, feasibility, safety, effectiveness, and tolerability of CAB+RPV-LA administration at NHS HIV clinics and community sites?
- What are the unmet needs of patients taking CAB+RPV-LA as part of their future routine clinical care?
- What are the characteristics and reasons of people living with HIV who are eligible but decline to take part in the ILANA trial?

## Methods and Analysis

### Study design

This is a 12-month, multi-centre mixed-methods study examining the implementation of CAB+RPV-LA injections in clinics and community-based settings in England in people who will receive CAB+RPV-LA as part of their routine care.

#### Study Setting

The study will take place at six NHS England HIV clinic sites: four in London (Barts Health NHS Trust, Chelsea and Westminster Hospital NHS Foundation Trust, Guy’s and St Thomas’ NHS Foundation Trust, Royal Free London NHS Foundation Trust), one in Liverpool and one in Brighton (Royal Liverpool University Hospital and Royal Sussex County Hospital).

The study has two phases (Figure 1):

1. **Phase 1 (Clinic Phase)** - implementation of CAB+RPV-LA in the clinic with evaluation and blueprint (Standard Operating Procedure) development.
2. **Phase 2 (Community/Hospital Phase)**- implementation of CAB+RPV-LA in the clinic and community setting with evaluation and blueprint (SOP) development.

**Figure 1.**
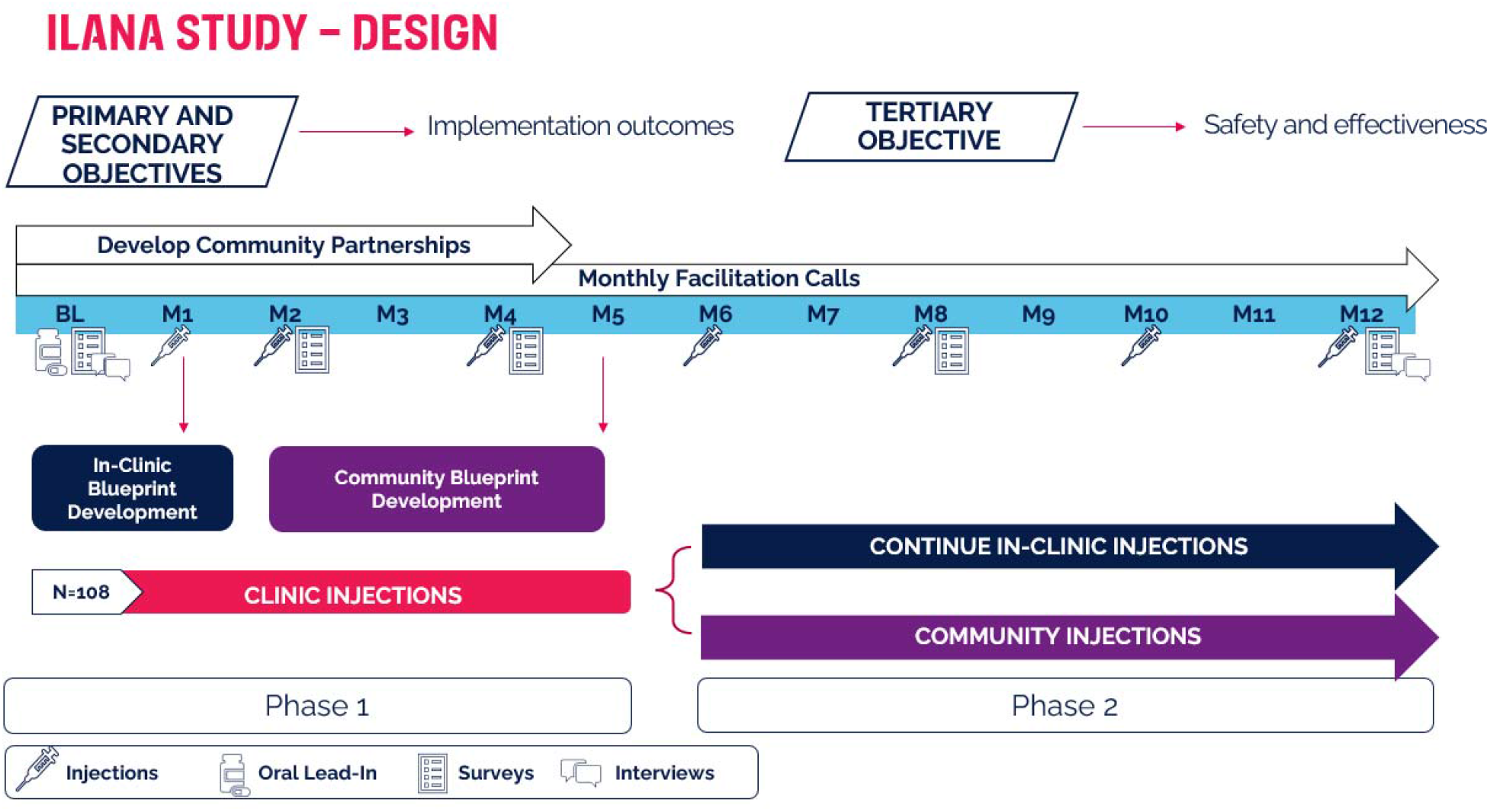
ILANA Study Design.

All participants begin injections in the clinic setting in Phase 1. During Phase 1, participants are screened and consented to participate. Potential participants will be asked to indicate their preferred location for Phase 2 (community or hospital site) and the reason for their preference. In Phase 2, participants will either continue to receive injections in clinic or at community sites (e.g. home, community organization, primary care). Each site has a target of 18-participants and agreed to adhere to recruitment caps to ensure recruitment of at least 50% women, 50% racially-minoritised people, 30% people over the age of 50 are enrolled.

### Outcome Measures

The study includes (i) participant outcomes; (ii) care provider and nurse participant outcomes; and (iii) community site representative outcomes (described in table 1).

**Table 1:** Outcome measures

| Participant outcomes |
| --- |
| **Primary outcome**<br>The primary outcome measure is the proportion of participants that agree or completely agree (average score of 4 or higher) on the Feasibility of Intervention Measure (**FIM**) at **12 months** in relation to the **injection**. |
| **Secondary outcomes**<br>The proportion of participants that agree or completely agree (average score of 4 or higher) on the **FIM** at **4 months** in relation to the **injection**.<br>The proportion of participants that agree or completely agree (average score of 4 or higher) on the **FIM** at **12 months** in relation to the **alternative community-based location**.<br>The proportion of participants that agree or completely agree (average score of 4 or higher) on the **FIM** at **4 months** in relation to the **alternative community-based location**.<br>Proportion participants that agree/completely agree (average score 4 or higher) on the |
| Acceptability of Intervention Measure (AIM) at **4 and 12 months** in relation to the **injection**<br>• Proportion participants that agree/completely agree (average score 4 or higher) on the **AIM** at **4 and 12 months** in relation to **the alternative community-based location**<br>• Change from baseline in HIV Treatment Satisfaction Questionnaire (**HIVTSQs-12**) at **4 and 12 months**<br>• Descriptive summaries of the tolerability and acceptance of injections as measured by the **acceptability/convenience of injections** questionnaire at **4 and 12 months**<br>• Change from baseline in Intervention Appropriateness Measure (**IAM**) score at **4 and 12 months** in relation to the **injection**<br>• Change from baseline in **IAM** score at **4 and 12 months** in relation to the **alternative community-based location**<br>• Descriptive summaries of the barriers and facilitators to implementation of CAB and RPV LA as measured by the questionnaires on **appointment scheduling, preferences for information, appointment reminders, appointment times and locations and stigma/fad concepts** measured at **4 and 12 months**<br>• Descriptive summaries for participant preferences for injection settings as measured by the questionnaires on **differentiating care settings, treatment location preferences and time spent in/getting to clinic** at **4 and 12 months** |
| **Adherence outcomes**<br>• Proportion participants who receive all injections on the target date up to 4 months from baseline<br>• Proportion participants who receive all injections on the target date up to 12 months from baseline<br>• Proportion participants who receive all injections within $\pm 7$ days of target date up to 4 months from baseline<br>• Proportion participants who receive all injections within $\pm 7$ days of target date up to 12 months from baseline<br>• Proportion participants who receive all injections within $\pm 14$ days of target date up to 4 months from baseline<br>• Proportion participants who receive all injections within $\pm 14$ days of target date up to 12 months from baseline<br>• Proportion of participant who missed at least one injection within 4 months and 12 months from baseline.<br>• Proportion participants who received oral bridging for all missed injections within 4 months and 12 months from baseline.<br>• Number of injections given outside 7-day window within 4 months and 12 months from baseline.<br>• Proportion of injections given outside 7 days where oral bridging was used within 4 months and 12 months from baseline.<br>• Number of participants with injections given outside 7-day window<br>• Proportion of participants with injections given outside 7 days where oral bridging was used |
| **Tertiary outcomes (relating to safety and clinical efficacy)** |
| • Proportion of participant with at least one and number of serious adverse event (SAE)<br>• Proportion of participants with at least one and number of adverse drug reactions (ADR) (excluding injection site reactions)<br>• Number of pregnancies and pregnancy outcomes<br>• Proportion of participants with at least one device failure and number of device failures<br>• Proportion of participants who fail to progress to injections/discontinuation during oral lead-in<br>• Proportion of participants who discontinue CAB and RPV LA<br>• Proportion who discontinue CAB and RPV LA due to SAE<br>• Proportion who discontinue CAB and RPV LA due to an adverse drug reaction (excluding injection site reactions)<br>• Proportion who discontinue CAB and RPV LA due to injection-related reason (injection site reactions or inconvenience)<br>• Proportion of participants within each of the following categories of HIV viral load: (VL) $\geq 200\text{c/ml}$, (VL) $\geq 50\text{c/ml}$ $<200\text{c/mL}$ or (VL) $< 50\text{c/ml}$ |
| **Provider outcomes: Care provider and nurse participant outcomes** |
| • The proportion of participants that agree or completely agree (average score of 4 or higher) on the Feasibility of Intervention Measure (FIM) at 4 and 12 months.<br>• Proportion of participants that agree/completely agree (average score 4 or higher) on the Acceptability of Intervention Measure (AIM) at 4 and 12 months<br>• Descriptive summaries of the barriers and facilitators to implementation of CAB and RPV LA at 4 and 12 months<br>• The proportion of participants that used the implementation Blueprint at 4 and 12 months |
| **Provider outcomes: Community site representative outcomes** |
| • The proportion of participants that agree or completely agree (average score of 4 or higher) on the Feasibility of Intervention Measure (FIM) at 8 and 12 months.<br>• Proportion participants that agree/completely agree (average score 4 or higher) on the Acceptability of Intervention Measure (AIM) at 8 and 12 months |

#### NO-LANA Sub-study

Those who decline to take part in the ILANA study are a group of individuals otherwise willing to commence on long-acting ART.T heir reasons for declining are unlikely to relate to concerns about starting a new regimen. This makes ILANA ‘non-participants’ an ideal group to explore the reasons why individuals do not wish to take part in research. A sub-study, called NO-LANA, will be carried out alongside ILANA in the same six study sites. It is a mixed-methods study based on an online survey and individual interviews with people living with HIV who declined to take part in ILANA.

##### Patient and public involvement and engagement

The need for the ILANA study emerged from clinical practice and discussions with service users, advocacy groups and healthcare professionals. Real word insights into the implementation of CAB+RPV-LA surfaced as a cross-cutting priority for the groups.

To create a platform to deliver the meaningful inclusion and authentic co-production of research, the research collaborative group (the SHARE Collaborative) have convened a paid community advisory board. Through an iterative process of consultation, review and implementation, the community advisory board are lending their living and lived experiences of HIV and their community expertise to this and other projects at every step, from study design to data collection and knowledge translation. A peer-researcher with lived experience of HIV is employed as an integral member of the research team and is a named collaborator, contributing to the development of the protocol and questionnaires ^(18)^, and will continue to contribute to delivery and meet ICJME criteria for authorship ^(19)^.

##### Recruitment

Sites were selected based on research experience and on willingness to agree to a proactive recruitment strategy with the application of caps. Weekly reporting is distributed via a newsletter to all sites with a detailed breakdown of age, gender and ethnicity of participants recruited and uses forecasting to monitor recruitment. If a site is unable to achieve the diversity targets, competitive recruitment will be applied. The peer researcher will ask local community groups to discuss the trial within their communities.

### Sample size

One hundred and eight virologically suppressed people living with HIV will be included in this study and will receive CAB+RPV-LA in the clinic and/or the community. There is no formal statistical comparison between groups. The NO-LANA sub-study has no set sample size as all participants who declined to take part in ILANA are eligible; it is estimated around 70 people will take part in NO-LANA.

### Participant identification

Potential participants who will be receiving CAB+RPV-LA within their routine clinical care will be referred to the study team by their clinic doctors within the HIV clinics of the six sites. The local HIV teams will inform participants of the trial and will provide patient information leaflets with contact details of the study team. Community organisations will inform their service users of the trial and trial sites. Local HIV teams will inform potential NO-LANA sub-study participants no sooner than one-month after they have been asked (and declined) to take part in ILANA that they are invited to take part in an anonymous survey and individual interviews.

### Inclusion and exclusion criteria

Adult people living with HIV (age≥18 years) will be invited to participate if they have capacity to consent with the following criteria:

- Will receive CAB+RPV-LA as part of their routine clinical care
- In accordance with UK SmPC and NICE guidance:
  - Virologically suppressed (HIV-1 RNA <50 copies/ml) on a stable antiretroviral regimen
  - Without present or past evidence of viral resistance to, and no prior virological failure with agents of the non-nucleoside reverse transcriptase inhibitor (NNRTI) and integrase inhibitor (INI) class
  - Not co-infected with Hepatitis B
  - Pregnancy – participants of child-bearing age will be advised as per routine clinical care that there are insufficient data to recommend the use of the drug in pregnancy but will not be expected to use contraception (ie as per SmPC license)

Participants who have previously taken NNRTIs will be assessed to ensure no previous viral resistance during the pre-screening process.

For the NO-LANA sub-study, anyone eligible for ILANA who declined the study is eligible.

#### Data collection

All participating sites will be assessed using a mixed-methods approach including questionnaires, semi-structured qualitative interviews, templated data collection instruments and primary data sources (clinic records). Clinical endpoints are tertiary and descriptive.

### Questionnaires

Questionnaires use established instruments and some tailored to the purpose of the study. Patient reported outcome measures such as the HIV treatment Satisfaction questionnaire will be used for participant data collection, and the AIM (Acceptability of Intervention Measure), IAM (Intervention Appropriateness Measure) and FIM (Feasibility of Intervention Measure) ^(20)^ will be adapted and anchored for implementation intervention. The NO-LANA sub-study questionnaire consists of a brief set of demographic questions and questions about reasons for non-participation in ILANA.

### Participant questionnaires

Questionnaires will be provided to participants at baseline, months one, four and twelve by clinicians involved in the study, during their scheduled clinic visit.

### Provider questionnaires

Community nurses and clinic nurses will complete questionnaires at baseline, and months four and twelve. Community representatives will complete questionnaires at months eight and twelve.

### In-depth semi-structured interviews

Non-probability purposive sampling will be used to recruit a diverse group of patients in terms of age, gender, socioeconomic status as participants in qualitative interviews.

Interviews aim to elicit narratives about specific experiences, fears, hopes, concerns and unexpected outcomes of utilizing CAB+RPV-LA. All interviews will be conducted by researchers trained in qualitative interview methodology and implementation science frameworks, guided by a semi-structured interview guide.

Interviews will be audio recorded and transcribed in full and are expected to last around 30 minutes.

Two interviews with people living with HIV will be conducted per clinic site at baseline and at M12. Interviews will cease when data ‘saturation’ is reached – this is commonly defined as the point at which collecting or observing more data will not lead to discovery of more information related to the research questions.

The study team will also approach two-four HCP participants (doctors, nurses, and non-clinical staff involved in the implementation and administration of CAB+RPV-LA) to take part in interviews, inform them about the objectives of the study, its benefits, and risks, and how long the interview will take. The interviews will take place at baseline and month twleve, with consent.

### NO-LANA sub-study

All those who were invited to be part in ILANA, and declined, will be asked whether they wish to take part in NO-LANA. If they agree, they will be sent a link to an online survey. The survey (the NO-LANA Survey) will be anonymous and include: study information and consent and data protection information; a set of questions on demographic characteristics; a set of questions asking for reasons why participants did not want to take part in ILANA.

On completion of the survey there will be an option to leave contact details to take part in an individual interview with a member of the research team to explore issues in more detail. Participants will receive an information sheet (with option of receiving this via the clinic, email or post) and will have the option of conducting the interview in the clinic, via phone, or video-call.

The one-to-one interviews will be semi-structured and designed to take around 30-mins to complete (although longer time will be allocated) in recognition of the evidence that time pressures might be amongst the reasons for non-participation. During the interview, participants will be asked to discuss, in the first instance, their main reasons for not taking part in the ILANA trial. This will be followed by a set of broader questions covering issues such as researchers’ communication, delivery, modality and asking participants for suggestions or preferences on how these might be improved in the future.

## Data analysis

### Qualitative analysis

Qualitative interviews will be audio-recorded, transcribed and coded. Thematic analysis will be adopted to give in-depth understanding of the data generated. Data will be transferred into NVivo for analysis using a mixture of inductive and deductive coding to explore and ‘plot’ reasons for non-participation. NO-LANA sub-study participants’ suggestions on improving recruitment strategies will be mapped and summarised in a set of recommendations alongside the ILANA research findings.

### Quantitative analysis

All participants who passed screening and entered the study (i.e.,completed baseline eCRF) will be included in the analysis population. Participants will be included in the analysis for each outcome if they (i) are part of the analysis population (defined above); and (ii) have recorded data for the outcome of interest (i.e., participants with missing outcome measures will be excluded). All data will be analysed using Stata version 17 or later (StataCorp, College Station. TX, USA). A detailed statistical analysis describing the full analyses is available in Appendix A.

### Participant flow and baseline characteristics

A Consolidated Standards of Reporting Trials (CONSORT) diagram will be constructed to describe participant flow through the ILANA study. Baseline characteristics will be summarised descriptively and will be presented by location (clinic or community) and overall. NO-LANA sub-study survey data will be analysed to produce descriptive statistics about demographic characteristics of non-participants.

### Analysis of primary and secondary outcomes

The analysis of the primary, secondary and tertiary (including safety) outcomes will be descriptive. Summary statistics will be provided for each outcome (mean (SD) or median (IQR), or number (percent)), depending on the type of outcome. For each outcome, we will summarise the number (%) of participants with available outcome data (the number included in the analysis, and conversely the number with missing outcome data and thus excluded from the analysis).

### Subgroup analyses

Subgroup analyses will be performed to investigate whether predefined outcome measures vary across healthcare setting (clinic/community) and participant characteristics: gender (man/woman/non-binary) and ethnicity (Black-African, Black-Caribbean, Black-British, Asian, White, Mixed Race, Other). Full details of the subgroup analyses to be performed can be found in appendix A.

## Ethics and dissemination

### Ethical considerations

All participants will be receiving CAB+RPV-LA as part of their routine care, regardless of the study. Any patient who does not wish to participate in this study, will still receive the treatment if they wish as part of their routine care.

Members of the healthcare team will seek verbal consent from potential participants to contact them with further study information. Once participants have verbally agreed to participate in an eligibility screening or to receive further information about the study, the informed consent process will begin. Prior to interviews, the researcher will again seek verbal consent. Ethical approval has been obtained from the Health Research Authority Research Ethics Committee (REC reference: 22/PR/0318).

### Data protection and patient confidentiality

Data protection has been central to the design. Data minimisation will be practised, with minimal personally identifiable information collected only when necessary. We will ensure an Information-governance tool-kit, GDPR and data-protection act compliant, robust, data-management policy and infrastructure. Data access will be restricted through a secure user-credentialing process.

Personal data collected for this project will include consent forms, medical data, questionnaires, recorded interviews, transcribed interviews and a digital key linking research participants to a unique code number (pseudoanonymisation code). This data will be transferred and stored securely within the QMUL data safe haven.

In the event that any hard copies of signed consent forms or of questionnaire are required, these will be stored in locked filing cabinets in an area with restricted access.

Interview audio files will be recorded on password-protected encrypted digital voice recorders and deleted from the from the digital voice recorder once directly uploaded onto encrypted hard drives. Audio-recordings will be assigned to the relevant unique study number and destroyed once they have been transcribed and checked. Hard copies of pseudonaymised transcripts will be stored in locked filing cabinets and on restricted-access, password-protected on QMUL safe haven.

For the NO-LANA sub-study survey, any contact detail provided to be contacted for further individual interviews will be collected in a separate survey page so details cannot be linked to the anonymous survey data.

## Output and dissemination

The ILANA dissemination strategy will be formulated in conjunction with the SHARE Collaborative Community Advisory Board and the study peer researcher to maximise the impact of this work on clinical care and policy. This strategy will draw upon and leverage existing resources within the participating organisations, such as their academic infrastructure, professional relationships and community networks. Results will be disseminated in a range of innovative and engaging ways, using study-specific Twitter and Instagram accounts and a study website, which allow active engagement with people living with HIV, HCPs, the academic community and the wider public.

## Discussion

Three seminal registrational trials demonstrate the clinical evidence of safety, effectiveness, tolerability of LAIs ARVs ^(6, 7, 21, 22)^. However, there is little published ‘real-world’ evidence and no published studies which focus on delivery in both the community and clinic settings, and none that focus on inclusion of women and racially-minoritised people. ^(23)^ Implementation science can reveal barriers, challenges, opportunities for real-world uptake and scale up of innovation, helping to bridge the gap between research theory and clinical practice. Our study will therefore contribute to translating knowledge by exploring patients’ and providers’ experiences and perceptions of LAIs ARVs to support implementation in pragmatic, real-world, settings in the clinic and community.

Although globally, 52% of people living with HIV are women, women, racially-minoritised and older people are chronically under-represented in HIV clinical trials ^(12, 13)^. A proactive recruitment cap in this study will ensure that we go beyond ‘lip-service’ and hold researchers to account when designing trials, including engaging with a peer researcher from the outset. Engaging actively with community organisations will support wider awareness of this implementation trial and improve the chances of LAIs benefitting those who might need it the most to support their adherence ^(24)^.

The reasons for minority underrepresentation in clinical research are varied and complex, hence longitudinal multi-phase qualitative research methods will be deployed to understand potential barriers for women and racially-minoritised people living with HIV. Engagement in care can be affected by unequal power dynamics between the healthcare provider and service-user, and there is a lack of research on inequities in implementation outcomes. Using an equity lens as the ILANA study will help identify and understand the specific and multiple dimensions affecting healthcare and medication engagement ^(25)^.

For example, research with Black-African communities in the UK found that patients’ concerns, influenced to varying degrees by migrant status in the UK, were important yet unappreciated factors shaping treatment adherence ^(26)^. The ILANA study explores participants’ perceptions and barriers such as social and economic hardship ^(27)^ which affect access to care, physical and mental health, and quality of life ^(28)^.

Exploring the use of alternative settings for injection, including community-based settings to deliver CAB+RPV-LA, has the potential to expand options for access whilst also potentially mitigating against the human resource requirements of delivering injections and improving capacity. Previous research has shown that community-based delivery of ART increases viral suppression among people living with HIV ^(29)^. Many people living with HIV report fear of encountering stigma when attending HIV clinics, which can affect engagement with care. Therefore, receiving care in a community setting may provide additional and preferable choices and the possibility of receiving treatment in a less medicalised setting ^(30)^. Shared decision-making between patient and healthcare providers builds trust and strengthen partnerships ^(31)^ which can support optimum transition from oral ART to injectable ART.

Education about LAIs for healthcare providers and people living with HIV is vital to the success of implementation. To facilitate increased availability of long-acting injectable options and ensure health equity for the UK and at global scale, it is vital to adapt healthcare systems and facilities and to develop pathways that work for a diverse range of people.

Finally, the NO-LANA sub-study was designed to capture the reasons for non-participation in ILANA, both as a form of reflexive practice and as an exemplar of factors that might drive reluctance to take part in research even when individuals are interested in the intervention on offer. Findings from NO-LANA will be applicable beyond the ILANA study through theoretical generalisation, illuminate barriers to research participation and gather patient-driven suggestions on how to address these.

## Limitations

The ILANA trial is primarily based in the NHS setting and in the Global North. The findings may not be reproducible in countries of the Global South where there remains an urgent need for LAIs but where health systems, epidemiological landscape and broader socio-economic contexts differ substantially from those in England.

Moreover, all ILANA sites are in large and inner-city settings, which leaves a data gap regarding implementation of LAIs in smaller and rural settings. The small sample size and specific targets for women and racially-minoritised groups aims at being representative of the HIV epidemic in the UK, but may not be representative of the population of patients choosing the injectable option.

Finally, the ILANA participants will have a history of good adherence which is needed to be eligible for LAIs in the NHS, which may mean the evidence of the impact of injectables on their adherence might prove less generalisable to wider populations living with HIV in the UK.

## Conclusion

Long-acting ART has the potential to move the field closer to the goal of offering ART that more people can accept and adhere to. With its novel study design, the ILANA trial aims to fulfill a significant knowledge gap about optimal implementation strategies with close attention to patient experience, real-world practice in health systems, and alternative access options for marginalized and underrepresented populations. Only by addressing these issues throughout our innovation efforts we can reduce health inequity for people living with HIV.

## Data Availability

All data produced in the present study are available upon reasonable request to the authors

## Supplementary Appendix

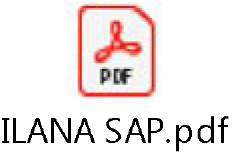

## References

1. Drake AL, Wagner A, Richardson B, John-Stewart G. Incident HIV during Pregnancy and Postpartum and Risk of Mother-to-Child HIV Transmission: A Systematic Review and Meta-Analysis. PLOS MEDICINE. 2014;11(2).

2. Ferrand RA, Corbett EL, Wood R, Hargrove J, Ndhlovu CE, Cowan FM, et al. AIDS among older children and adolescents in Southern Africa: projecting the time course and magnitude of the epidemic. AIDS. 2009;23(15):2039–46.

3. UN/WHO. UNAIDS data 2018 - UN Joint Programme on HIV/AIDS (UNAIDS) 2018 [Available from: http://www.unaids.org/en/resources/documents/2018/unaids-data-2018.

4. Ford N, Migone C, Calmy A, Kerschberger B, Kanters S, Nsanzimana S, et al. Benefits and risks of rapid initiation of antiretroviral therapy. AIDS. 2018;32(1).

5. Swindells S, Lutz T, Van Zyl L, Porteiro N, Stoll M, Mitha E, et al. Week 96 extension results of a Phase 3 study evaluating long-acting cabotegravir with rilpivirine for HIV-1 treatment. AIDS. 2022;36(2).

6. Orkin C, Oka S, Philibert P, Brinson C, Bassa AC, Gusev D, et al. Long-acting cabotegravir + rilpivirine for HIV treatment: FLAIR week 96 results. HIV Medicine. 2020;21(SUPPL 4):14.

7. Orkin C, Oka S, Philibert P, Brinson C, Bassa A, Gusev D, et al. Long-acting cabotegravir plus rilpivirine for treatment in adults with HIV-1 infection: 96-week results of the randomised, open-label, phase 3 FLAIR study. LANCET HIV. 2021;8(4):e185–e96.

8. Chounta V, Overton ET, Mills A, Swindells S, Benn PD, Vanveggel S, et al. Patient-Reported Outcomes Through 1 Year of an HIV-1 Clinical Trial Evaluating Long-Acting Cabotegravir and Rilpivirine Administered Every 4 or 8 Weeks (ATLAS-2M). Patient. 2021;14(6):849–62.

9. Flamm J, Garris C, D’Amico R, Dalessandro M, McHorney CA, Mansukhani SG, et al. PB4 Patient Perspectives on Implementation of a Long-Acting Injectable Antiretroviral Therapy Regimen in HIV US Healthcare Settings: Final Month 12 Results from the CUSTOMIZE Study. Value in Health. 2021;24:S13.

10. Havlir D, Gandhi M. Implementation challenges for long-acting antivirals as treatment. Current Opinion in HIV and AIDS. 2015;10(4).

11. Hedge B, Devan K, Catalan J, Cheshire A, Ridge D. HIV-related stigma in the UK then and now: to what extent are we on track to eliminate stigma? A qualitative investigation. BMC Public Health. 2021;21(1):1022.

12. Castillo-Mancilla JR, Cohn SE, Krishnan S, Cespedes M, Floris-Moore M, Schulte G, et al. Minorities remain underrepresented in HIV/AIDS research despite access to clinical trials. HIV clinical trials. 2014;15(1):14–26.

13. Curno MJ, Rossi S, Hodges-Mameletzis I, Johnston R, Price MA, Heidari S. A Systematic Review of the Inclusion (or Exclusion) of Women in HIV Research: From Clinical Studies of Antiretrovirals and Vaccines to Cure Strategies. JAIDS Journal of Acquired Immune Deficiency Syndromes. 2016;71(2).

14. Dhairyawan R, Okhai H, Hill T, Sabin CA. Differences in HIV clinical outcomes amongst heterosexuals in the United Kingdom by ethnicity. AIDS. 2021;35(11).

15. UNAIDS. 2020 Global AIDS update—seizing the moment—tackling entrenched inequalities to end epidemics.

16. O’Halloran C, Sun S, Nash S, Brown A, Croxford S N C. HIV in the United Kingdom: Towards Zero 2030. 2019 report. London: Public Health England; 2019.

17. Orkin C, Apea V. International Women’s Day—how can I help? The Lancet HIV. 2022;9(4):e228–e9.

18. Devotta K, Woodhall-Melnik J, Pedersen C, Wendaferew A, Dowbor TP, Guilcher SJT, et al. Enriching qualitative research by engaging peer interviewers: a case study. Qualitative Research. 2016;16(6):661–80.

19. ICMJE. Defining the Role of Authors and Contributors 2021 [Available from: http://www.icmje.org/recommendations/browse/roles-and-responsibilities/defining-the-role-of-authors-and-contributors.html.

20. Weiner BJ, Lewis CC, Stanick C, Powell BJ, Dorsey CN, Clary AS, et al. Psychometric assessment of three newly developed implementation outcome measures. Implementation Science. 2017;12(1):108.

21. Orkin C, Arasteh K, Hernandez-Mora MG, Pokrovsky V, Overton ET, Girard P-M, et al. Long-acting cabotegravir + rilpivirine for HIV maintenance: Flair week 48 results. Topics in Antiviral Medicine. 2019;27(SUPPL 1):52s–3s.

22. Orkin C, Arasteh K, Górgolas Hernández-Mora M, Pokrovsky V, Overton ET, Girard P-M, et al. Long-Acting Cabotegravir and Rilpivirine after Oral Induction for HIV-1 Infection. New England Journal of Medicine. 2020;382(12):1124–35.

23. Schlechter CR, Del Fiol G, Lam CY, Fernandez ME, Greene T, Yack M, et al. Application of community – engaged dissemination and implementation science to improve health equity. Preventive Medicine Reports. 2021;24:101620.

24. Glandon D, Paina L, Alonge O, Peters DH, Bennett S. 10 Best resources for community engagement in implementation research. Health Policy and Planning. 2017;32(10):1457–65.

25. Baumann AA, Cabassa LJ. Reframing implementation science to address inequities in healthcare delivery. BMC Health Services Research. 2020;20(1):190.

26. Thomas F, Aggleton P, Anderson J. ‘Experts’, ‘partners’ and ‘fools’: Exploring agency in HIV treatment seeking among African migrants in London. Social Science & Medicine. 2010;70(5):736–43.

27. Ibrahim F, Anderson J, Bukutu C, Elford J. Social and economic hardship among people living with HIV in London. HIV medicine. 2008;9:616–24.

28. Solomon D, Tariq S, Alldis J, Burns F, Gilson R, Sabin C, et al. Ethnic inequalities in mental health and socioeconomic status among older women living with HIV: results from the PRIME Study. Sexually Transmitted Infections. 2022;98(2):128.

29. Barnabas RV, Szpiro AA, van Rooyen H, Asiimwe S, Pillay D, Ware NC, et al. Community-based antiretroviral therapy versus standard clinic-based services for HIV in South Africa and Uganda (DO ART): a randomised trial. The Lancet Global Health. 2020;8(10):e1305–e15.

30. Zheng C, Wang W, Young SD. Identifying HIV-related digital social influencers using an iterative deep learning approach. AIDS. 2021;35:S85–S9.

31. Kanazawa JT, Saberi P, Sauceda JA, Dubé K. The LAIs Are Coming! Implementation Science Considerations for Long-Acting Injectable Antiretroviral Therapy in the United States: A Scoping Review. AIDS Research and Human Retroviruses. 2020;37(2):75–88.

